# Eligibility for Subcutaneous Implantable Cardioverter Defibrillator in Congenital Heart Disease

**DOI:** 10.1101/19009175

**Authors:** Linda Wang, Neeraj Javadekar, Ananya Rajagopalan, Nichole M. Rogovoy, Kazi T. Haq, Craig S. Broberg, Larisa G. Tereshchenko

**Affiliations:** Oregon Health & Science University, Knight Cardiovascular Institute, Portland, OR

**Keywords:** subcutaneous ICD, electrocardiogram, eligibility

## Abstract

**Background:** The goals of this study were: assess left-and right-sided subcutaneous implantable cardioverter-defibrillator (S-ICD) eligibility in adult congenital heart disease (ACHD) patients, use machine learning to predict S-ICD eligibility in ACHD patients, and transform 12-lead ECG to S-ICD 3-lead ECG, and vice versa.

**Methods:** ACHD outpatients (n=101; age 42±14 y; 52% female; 85% white; left ventricular ejection fraction (LVEF) 56±9%) were enrolled in a prospective study. Supine and standing 12-lead ECG was recorded simultaneously with a right- and left-sided S-ICD 3-lead ECG. Peak-to-peak QRS and T amplitudes, RR, PR, QT, QTc, QRS intervals, T_max_, and R/T_max_ (31 predictor variables) were tested. Model selection, training, and testing were performed using supine ECG datasets. Validation was performed using standing ECG datasets and out-of-sample non-ACHD population (n=68; age 54±16 y; 54% female; 94% white; LVEF 61±8%).

**Results:** A 40% of participants were ineligible for S-ICD. Tetralogy of Fallot patients passed right-sided screening (57%) more often than left-sided (21%; McNemar’s χ^2^ *P*=0.025). The ridge model demonstrated the best cross-validation function. Validation of the ridge models was satisfactory for standing left-sided [ROC AUC 0.687 (95%CI 0.582-0.791)] and right-sided [ROC AUC 0.655(95%CI 0.549-0.762)] S-ICD eligibility prediction. Out-of-sample validation in the non-ACHD population yielded a 100% sensitivity of the pre-selected threshold for the elastic net model. Validation of the transformation matrices showed satisfactory agreement (<0.1 mV difference).

**Conclusion:** Nearly half of the contemporary ACHD population is ineligible for S-ICD. Machine-learning prediction of S-ICD eligibility can be used for screening of S-ICD candidates.

## Introduction

A subcutaneous implantable cardioverter-defibrillator (S-ICD) is a life-saving device that prevents sudden cardiac arrest in vulnerable patients.^1^ The approval of the S-ICD for use in the United States is significant because of the benefits it has over the traditional, transvenous ICD, which include the lack of risk for vascular occlusion, systemic infection^2^, and the adverse effects of lead extraction.^3^ S-ICD can be especially advantageous in adults with congenital heart disease (ACHD) patients who may have limited or no venous access to the heart, and in whom there is increased risk of systemic embolism when a persistent shunt is present.^4, 5^ These individuals are often at increased risk for sudden cardiac arrest that is higher in ACHD compared to the general population^6^ and frequently require thoracic surgery to place an epicardial ICD system. ACHD patients may face multiple generator changes in their lifetime, making an S-ICD a viable option due to its less-invasive placement. The 2017 AHA/ACC/HRS Guidelines^7^ for the prevention of sudden cardiac arrest in ACHD patients recommend S-ICD use when feasible.

S-ICD requires electrocardiographic (ECG) pre-screening before implantation to assess sensing. S-ICD screening involves the recording of a special 3-lead ECG with ECG electrodes placed in the locations of S-ICD sensing electrodes, as advised by the manufacturer.^8^ This additional step may negatively impact the utilization of S-ICD.^9^ Lack of confidence is the most common barrier for referral^10^ among physicians, and the perceived strength of the physician recommendation is the most common theme associated with ICD refusal among primary prevention candidates.^11^ Conversely, a 12-lead ECG is readily available and easy to obtain. Therefore, using a conventional 12-lead ECG as the tool for pre-screening eligibility would greatly improve a physician’s confidence and recommendation to suitable patients.

Our group recently developed a screening tool to predict left-sided S-ICD eligibility from a 12-lead ECG.^12^ However, validation of this screening tool in an out-of-sample population has not been performed. Moreover, in ACHD patients, right-sided S-ICD implantation may improve S-ICD eligibility.^13^ However, very little data is available regarding right-sided S-ICD eligibility predictors in ACHD patients. Furthermore, it remains unknown whether it is feasible to transform the 12-lead ECG into left-and right-sided S-ICD 3-lead ECG, and vice versa.

We conducted this study with several goals: (1) assess left-and right-sided S-ICD eligibility in ACHD patients, (2) validate the previous^12^ left-sided S-ICD eligibility prediction tool, (3) use machine learning to predict right- and left-sided S-ICD eligibility in ACHD patients, and (4) develop and validate transformation matrices to transform 12-lead ECG to S-ICD 3-lead ECG, and vice versa.

## Methods

A MATLAB (MathWorks, Inc, Natick, MA) open-source code for ECG analyses and a user manual is provided at https://github.com/Tereshchenkolab/S-ICD_eligibility. Fully de-identified raw digital ECG signal data generated for this study are available at the GitHub at https://github.com/Tereshchenkolab/S-ICD_eligibility.

### Study population

We conducted a prospective cross-sectional study at the Oregon Health & Science University (OHSU). The Institutional Review Board approved the study, and all participants signed informed consent before entering the study. Eligible adult patients that had been previously diagnosed with ACHD were invited to participate during scheduled appointment with their cardiologist. Inclusion criteria were: (1) known congenital heart defect followed at the OHSU ACHD clinic, (2) age ≥ 18 years, (3) able to stand on their own for the duration of ECG recording. Exclusion criteria were: (1) acute medical condition, (2) life expectancy less than one year due to non-cardiac condition and (3) developmental delay.

Study participants were grouped based on the complexity of ACHD anatomy and physiology as described in 2019 ACHD AP Classification^14^: simple (IA-B), moderate complexity (IIB-C), or complex(IIIC-D).

For out-of-sample validation of the machine learning models, we used the data of our previous S-ICD eligibility study,^12^ which enrolled a widely generalizable sample of the OHSU outpatient population, with a broad range of age (18–81 y), body mass index (BMI; 19–53 kg/m^2^), QRS duration (66–150 ms), and left ventricular ejection fraction (LVEF; 37–77%).

### ECG recording and traditional ECG analysis

A MAC 5500 HD ECG system (General Electric (GE) Healthcare, Milwaukee, WI, USA) was used to record ECGs. Four 10-second digital ECGs (sampling rate 500 Hz, amplitude resolution 1 µV) were recorded in the following order: right-sided 15-lead supine, left-sided 15-lead supine, left-sided 15-lead standing, and right-sided 15-lead standing. A15-lead ECG configuration included simultaneously recoded standard 12-lead ECG, and a special 3-lead ECG. Three additional unipolar ECG electrodes (*a*1, *a*2, *a*3) were placed according to recommendations^8^ to imitate the location of the sensing S-ICD electrodes (Figure 1).

**Figure 1.**
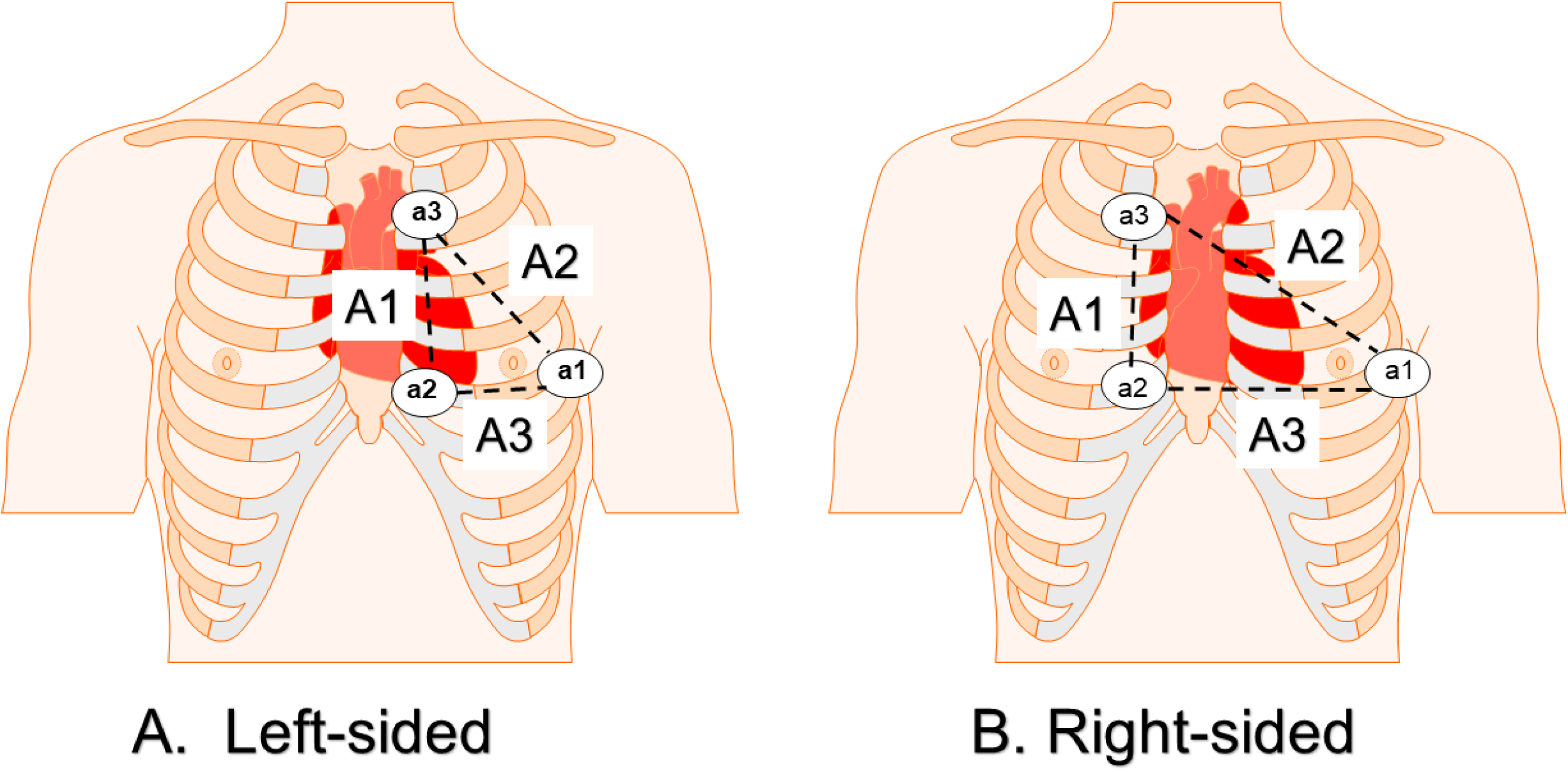
Placement of *a*1, *a*2, and *a*3 electrodes for the 3-lead ECG to mimic the leads A1 (*a*2-*a*3), A2 (*a*1-*a*3), and A3 (*a*1-*a*2) sensing vectors of the S-ICD.

For left-sided S-ICD, the *a*1 electrode was placed over the 5^th^ intercostal space at the midaxillary line, the *a*2 was placed 1 cm left lateral of the xiphoid midline, and the *a*3 was placed 14 cm superior to the *a*2 electrode.^8^ For right-sided S-ICD, *a*2 was moved 1 cm right lateral of the xiphoid midline, and *a*3 to 14 cm superior to the new right-sided placement of *a*2, whereas the position of *a*1 was left unchanged (Figure 1). In patients with dextrocardia, all electrodes were placed in mirror-image fashion, as appropriate.

Averaged across 12-leads RR’, PR, QT, QTc, and QRS intervals, as well as lead-specific peak-to-peak QRS- and T-amplitudes were measured automatically by the GE 12SL algorithm (GE Marquette, Milwaukee, WI), using Magellan ECG Research Workstation V2 (GE Marquette, Milwaukee, WI). R/T ratio in the lead with the largest T wave was calculated as previously described.^12^

### Anthropometric measurements

Hip, waist, lower and upper chest circumference were measured using a measuring tape while the participant was standing. The lower chest circumference was measured at the level of the xiphoid process, and upper chest circumference was measured at the level of the armpits. The subcostal angle was assessed. The ratio of the lateral diameter of the chest to the anteroposterior diameter of the chest was estimated. Height and weight were measured, and BMI was calculated.

### Assessment of S-ICD eligibility

Bipolar S-ICD leads were derived from recorded unipolar *a*1, *a*2, and *a*3 leads by subtraction, as follows: Bipolar lead A1 = *a*2 – *a*3. Bipolar lead A2 = *a*1 – *a*3. Bipolar lead A3 = *a*1 – *a*2. Digital bipolar 3-lead left- and right-sided ECG morphologies in standing and supine position were evaluated using a digitized version of the Boston Scientific EMBLEM S-ICD Patient Screening tool^8^ by at least two investigators (AR, NJ, LW). A MATLAB (Mathworks, Natick, MA) viewer for digital S-ICD eligibility assessment (Supplemental Figure 1) was developed by the investigators (NJ, KTH, AR). We provided open-source code and a user manual at https://github.com/Tereshchenkolab/S-ICD_eligibility. In the case of disagreement, the 3^rd^ investigator (LGT) made the final determination. A sensing vector passed screening if maximum QRS amplitudes crossed the dotted line and all QRS complexes and T waves fit within a profile in all beats, in both standing and supine 10-second recording at 5–20 mm/mV gain, either on the left or right side. The reasons for failure (high T-wave, high R-wave, deep S-wave, small QRS complex, high P, or flutter F-wave) were recorded.

### Statistical analyses

After confirmation of normality, continuous variables were reported as mean ± standard deviation (SD) and compared using the *t*-test. The χ^2^ test was used to compare categorical variables. A paired *t*-testing was used to compare ECG measurements on the left and right side, standing, and supine. McNemar’s χ^2^ statistic was used for paired comparison of S-ICD ineligibility causes in different positions (standing, supine) on the left and right side.

### Validation of the previous left-sided S-ICD eligibility tool

Accuracy of our previously developed left-sided S-ICD eligibility prediction tool^12^ was validated using the entire study population. We measured Area Under the Receiver Operating Characteristic Curve (ROC AUC), and calculated sensitivity and specificity of the previously defined threshold (pass if ≥ 0).

### Machine learning model selection, training, testing, and validation

We applied a machine learning technique to develop a prediction of left-sided and right-sided S-ICD eligibility. Supine ECG datasets served for machine learning (training and testing), whereas standing ECG datasets, and the data of our previous S-ICD eligibility study^12^ served for validation. We compared logistic regression, lasso, elastic net, and ridge models in four machine learning steps (Figure 2).

**Figure 2.**
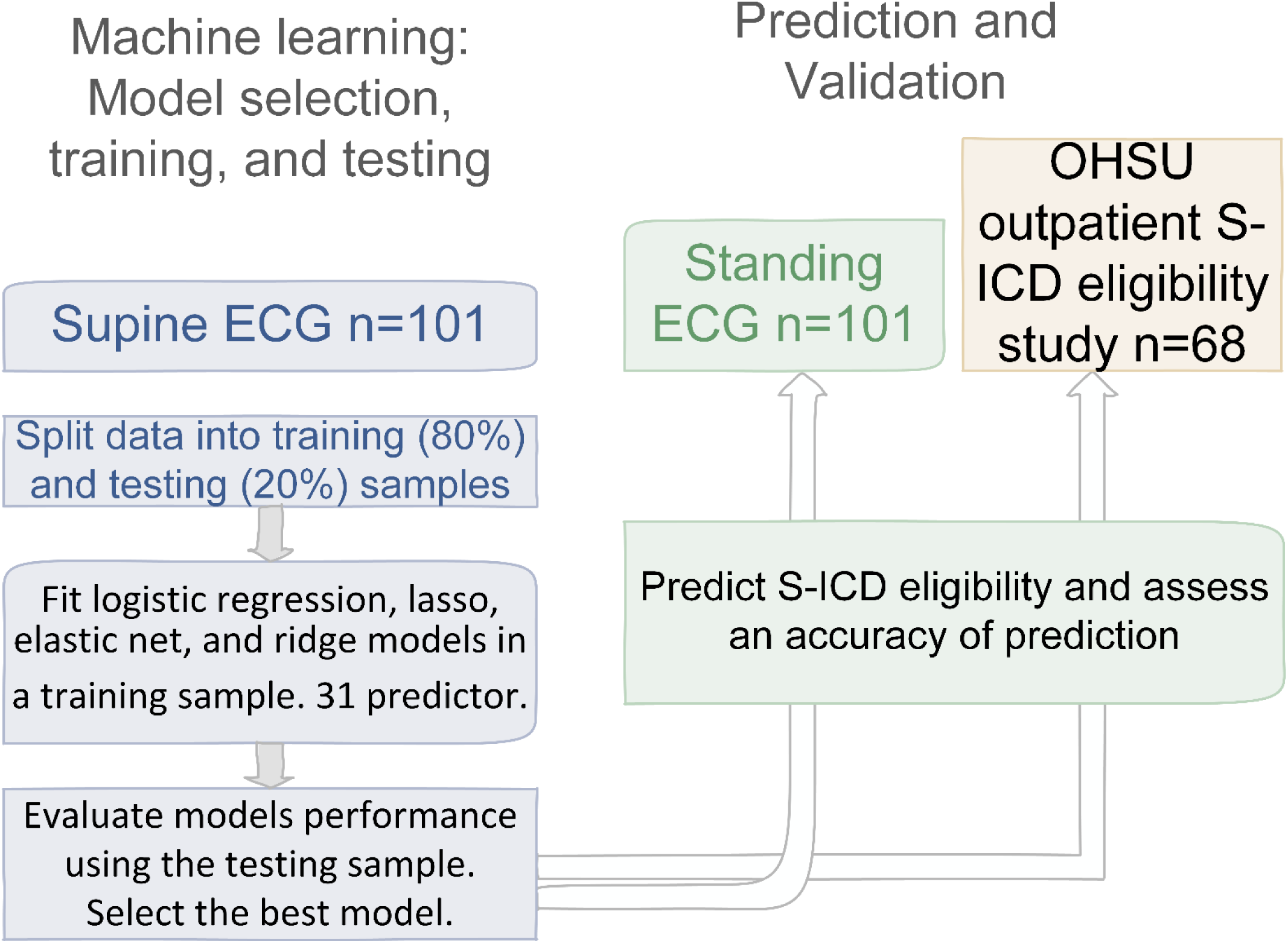
Machine learning steps: S-ICD eligibility prediction development, and validation.

In the first step, we split the data of the *supine* ECG datasets into training (80%) and testing (20%) random samples.

In the second step, we fitted four different models (logistic regression, lasso, elastic net, and ridge) using the training sample only. We included altogether 31 predictor variables: peak-to-peak amplitudes of QRS complex, T-amplitudes on each out of 12 ECG leads, and averaged across 12 leads PR, QT, QTc, QRS intervals, the largest T wave amplitude (T_max_), R/T_max_ ^15^, and heart rate (HR).

In the third step, we evaluated the prediction model performance of each technique (logistic regression, lasso, elastic net, and ridge) using the testing sample. We selected the best model with the smallest out-of-sample deviance and the largest deviance ratio. Penalized coefficients were used for comparison. The best threshold of predicted probability of left-sided and right-sided S-ICD eligibility in both training and testing samples was selected considering two factors. First, we considered the Liu method, which maximizes the product of sensitivity and specificity.^16^ However, as the goal of screening is to identify all individuals who are likely eligible for S-ICD, we strived to maximize the sensitivity of the test, targeting 100% sensitivity.

In the fourth step, we predicted left-sided and right-sided S-ICD eligibility in two new datasets: (1) *standing* ECG recordings, and (2) our previous S-ICD eligibility study data.^12^ We determined the accuracy of prediction by measuring ROC AUC. In addition, we validated the sensitivity of pre-defined (determined at the 3^rd^ step) threshold.

### Transformation of 12-lead ECG into S-ICD 3-lead ECG

The dataset was randomly split into the two equal size samples: the training and the validation samples each had 50% of the observations. Transformation matrices were developed for right-sided and left-sided, supine and standing sets of ECG data, to transform 12-lead ECG signal into 3-lead ECG signal, using random effect panel data linear regression with maximum likelihood estimator. Inverse transformation matrices were developed for transformation of S-ICD 3-lead ECG signal into 12-lead ECG signal. Previously, Kors et al. demonstrated the superiority of a statistical regression approach for the development of a transformation matrix.^17^ Transformation matrices were developed in the training sample. Then, in the validation sample, an agreement between the originally recorded and transformed 10-second signal was measured sample-by-sample, by paired *t*-testing, and the average difference with 95% confidence interval (CI) was reported.

Statistical analysis was performed using STATA MP 16.0 (StataCorp LP, College Station, TX). P-value < 0.05 was considered statistically significant.

## Results

### Study population

A total of 101 ACHD patients were recruited (Table 1). Most of the study participants had moderate or complex ACHD with hemodynamic impairment and on average, borderline systemic ventricular function. Participants had a history of Fontan, Ross, Mustard, Senning, Rastelli, Glenn, Damus-Kaye-Sensel, and Norwood procedures. Nearly every fifth study participant already had a transvenous cardiac device implanted: more likely an ICD (65%) than a pacemaker (35%). Approximately two-thirds of participants (68%) were currently taking cardiovascular medications (Table 1), and nearly half were taking antiarrhythmic medications (beta-blockers, calcium channel blockers, sotalol, amiodarone, dofetilide, or digoxin). Almost half of the study population was on anticoagulants or antiplatelet drugs. More than half of the population received drugs targeting hemodynamic improvement (angiotensin-converting enzyme inhibitors (ACEi), angiotensin receptor blockers (ARBs), angiotensin receptor-neprilysin inhibitor, aldosterone antagonists, vasodilators, and diuretics).

**Table 1.**
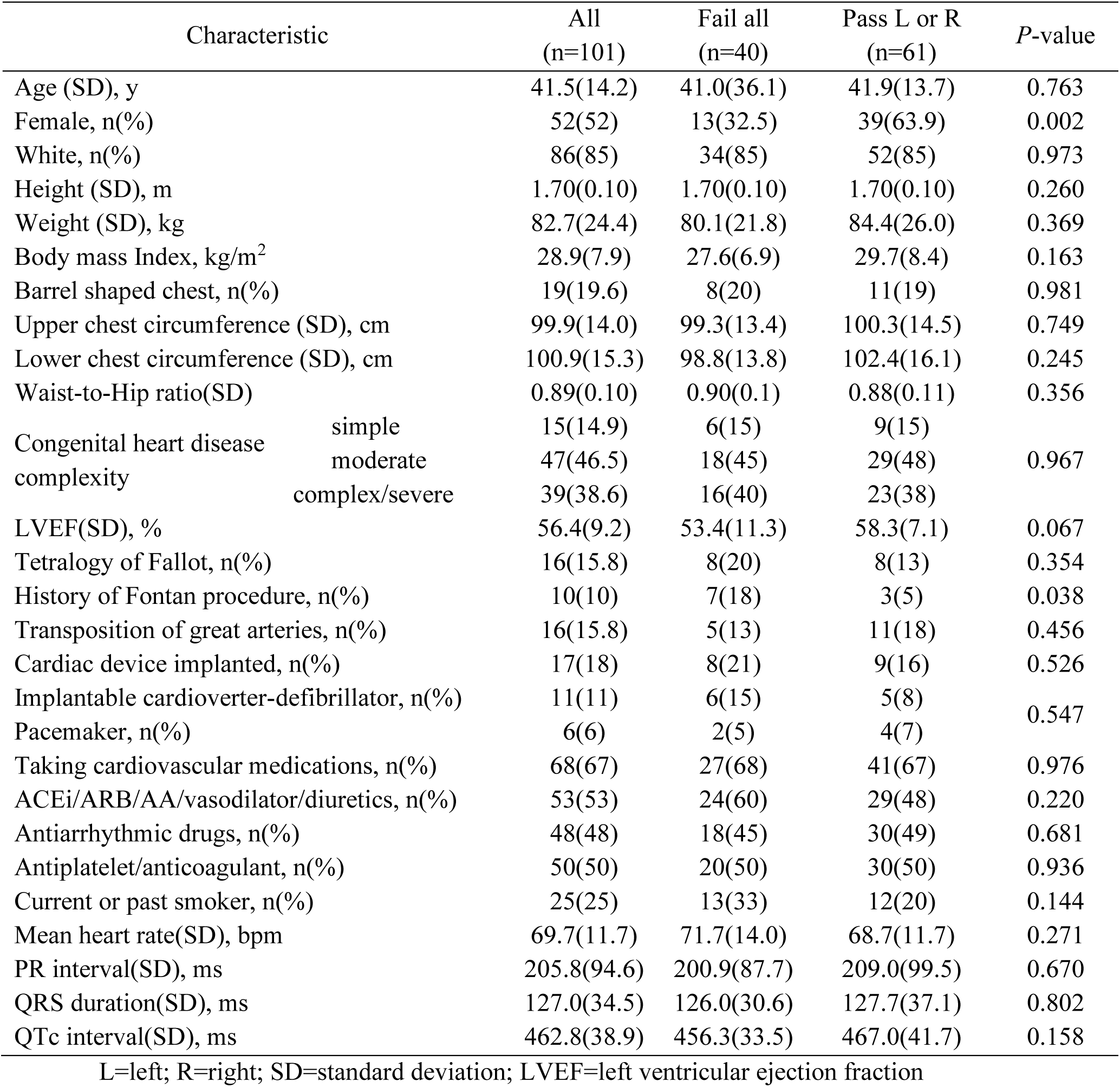
Clinical characteristics of the study participants

| Characteristic |  | All<br>(n=101) | Fail all<br>(n=40) | Pass L or R<br>(n=61) | P-value |
| --- | --- | --- | --- | --- | --- |
| Age (SD), y |  | 41.5(14.2) | 41.0(36.1) | 41.9(13.7) | 0.763 |
| Female, n(%) |  | 52(52) | 13(32.5) | 39(63.9) | 0.002 |
| White, n(%) |  | 86(85) | 34(85) | 52(85) | 0.973 |
| Height (SD), m |  | 1.70(0.10) | 1.70(0.10) | 1.70(0.10) | 0.260 |
| Weight (SD), kg |  | 82.7(24.4) | 80.1(21.8) | 84.4(26.0) | 0.369 |
| Body mass Index, kg/m<sup>2</sup> |  | 28.9(7.9) | 27.6(6.9) | 29.7(8.4) | 0.163 |
| Barrel shaped chest, n(%) |  | 19(19.6) | 8(20) | 11(19) | 0.981 |
| Upper chest circumference (SD), cm |  | 99.9(14.0) | 99.3(13.4) | 100.3(14.5) | 0.749 |
| Lower chest circumference (SD), cm |  | 100.9(15.3) | 98.8(13.8) | 102.4(16.1) | 0.245 |
| Waist-to-Hip ratio(SD) |  | 0.89(0.10) | 0.90(0.1) | 0.88(0.11) | 0.356 |
| Congenital heart disease<br>complexity | simple | 15(14.9) | 6(15) | 9(15) | 0.967 |
|  | moderate | 47(46.5) | 18(45) | 29(48) |  |
|  | complex/severe | 39(38.6) | 16(40) | 23(38) |  |
| LVEF(SD), % |  | 56.4(9.2) | 53.4(11.3) | 58.3(7.1) | 0.067 |
| Tetralogy of Fallot, n(%) |  | 16(15.8) | 8(20) | 8(13) | 0.354 |
| History of Fontan procedure, n(%) |  | 10(10) | 7(18) | 3(5) | 0.038 |
| Transposition of great arteries, n(%) |  | 16(15.8) | 5(13) | 11(18) | 0.456 |
| Cardiac device implanted, n(%) |  | 17(18) | 8(21) | 9(16) | 0.526 |
| Implantable cardioverter-defibrillator, n(%) |  | 11(11) | 6(15) | 5(8) | 0.547 |
| Pacemaker, n(%) |  | 6(6) | 2(5) | 4(7) |  |
| Taking cardiovascular medications, n(%) |  | 68(67) | 27(68) | 41(67) | 0.976 |
| ACEi/ARB/AA/vasodilator/diuretics, n(%) |  | 53(53) | 24(60) | 29(48) | 0.220 |
| Antiarrhythmic drugs, n(%) |  | 48(48) | 18(45) | 30(49) | 0.681 |
| Antiplatelet/anticoagulant, n(%) |  | 50(50) | 20(50) | 30(50) | 0.936 |
| Current or past smoker, n(%) |  | 25(25) | 13(33) | 12(20) | 0.144 |
| Mean heart rate(SD), bpm |  | 69.7(11.7) | 71.7(14.0) | 68.7(11.7) | 0.271 |
| PR interval(SD), ms |  | 205.8(94.6) | 200.9(87.7) | 209.0(99.5) | 0.670 |
| QRS duration(SD), ms |  | 127.0(34.5) | 126.0(30.6) | 127.7(37.1) | 0.802 |
| QTc interval(SD), ms |  | 462.8(38.9) | 456.3(33.5) | 467.0(41.7) | 0.158 |

L=left; R=right; SD=standard deviation; LVEF=left ventricular ejection fraction

### Assessment of S-ICD eligibility

There were 61 participants (60%) that passed either left- or right-sided screening, whereas the remainder of participants (40%) were deemed non-eligible for S-ICD. Ineligible participants were more likely to be males, or have had a Fontan palliation. There was a trend towards lower LVEF, the use of medications for heart failure treatment, history of past or current smoking, and lower BMI in those who failed ECG screening (Table 1). No difference in ACHD complexity was observed between those who passed versus failed screening.

Overall, a similar percentage of participants was eligible for right-sided (n=49; 49%) and left-sided S-ICD (n=45; 45%; McNemar’s χ^2^ *P*=0.450). Only a third of participants (n=33; 33%) passed both left- and right-sided screening, whereas 12 (12%) passed only left-sided, and 16 (16%) passed only right-sided screening. Tetralogy of Fallot patients passed right-sided screening (8/16) more often than left-sided (3/16; McNemar’s χ^2^ *P*=0.025). Similarly, taken together Tetralogy of Fallot and Fontan procedure patients (Figure 3) passed right-sided screening more often than left-sided (McNemar’s χ^2^ *P*=0.014). No anthropometric characteristics were associated with differences in either left- or right-sided S-ICD eligibility.

**Figure 3.**
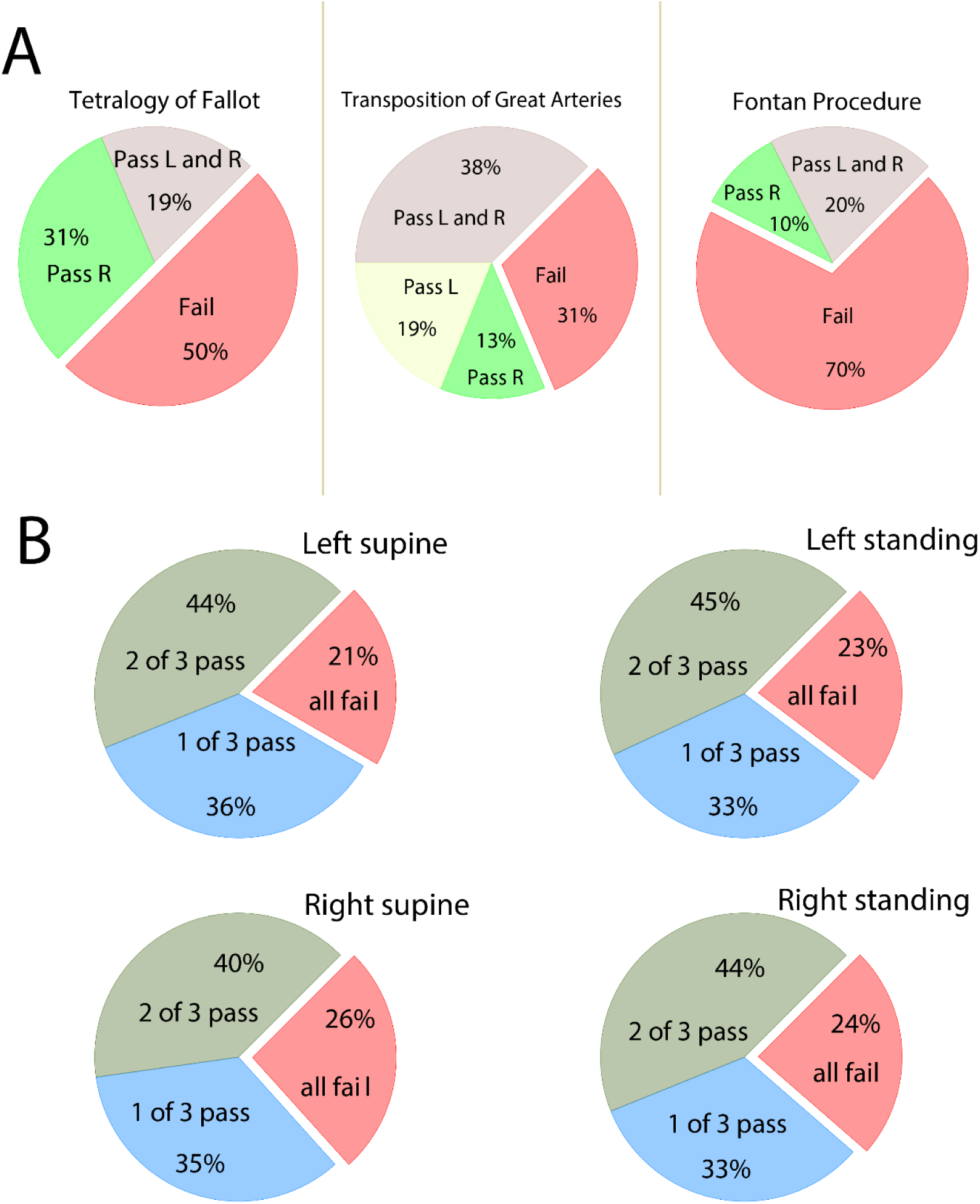
**A**. The proportion of patients with transposition of great arteries, Tetralogy of Fallot, and Fontan procedure with passing and failing for right (R)- and left (L)-sided sensing vectors. **B**. The proportion of study participants who failed all three vectors or passed 1-2 left- and right-sided vectors standing and supine.

No participants had all 3 S-ICD vectors with eligible ECG morphologies. In any position and any side, less than half of the participants (40-45%) had two admissible S-ICD vectors, whereas nearly a quarter of participants failed all three vectors (Figure 3).

Overall, little difference was observed in eligibility of ECG morphologies in different positions: left and right supine, left and right standing. The rates of pass/fail across complexity groups were similar for both right and left-sided vectors, either standing or supine (Figure 3). Representative examples of failed ECG morphologies are shown (Figure 4). Change of the body position from supine to standing led to a slight heart rate increase, QTc lengthening, and QRS shortening (Table 2). S-ICD ineligibility due to large P (or F) waves was more likely in left standing than in left supine position. S-ICD ineligibility due to a small QRS was more likely on the right side, in both supine and standing positions (Table 2). No other differences in ECG morphology affected S-ICD eligibility in different positions and sides.

**Table 2.** Comparison of ECG measurement and causes of S-ICD ineligibility for left- and right-sided ECG, standing and supine.

| Causes | Supine<br>Left-sided | Standing<br>Left-sided | *P Left Sup-*<br>*stand* | Supine<br>Right-sided | Standing<br>Right-sided | *P Right*<br>*sup-stand* | *P L-R*<br>*supine* | *P L-R*<br>*standing* |
| --- | --- | --- | --- | --- | --- | --- | --- | --- |
| Mean heart rate(SD), bpm | 70.0(12.7) | 76.1(13.6) | **<0.0001** | 70.4(13.3) | 76.1(13.2) | **<0.0001** | 0.070 | 0.687 |
| Mean QTc(SD), ms | 463.3(38.8) | 471.5(40.6) | **<0.0001** | 461.5(40.7) | 471.3(39.2) | **<0.0001** | 0.517 | 0.920 |
| Mean QRS(SD), ms | 127.1(34.7) | 120.5(34.7) | **<0.0001** | 127.0(34.4) | 122.7(34.5) | **0.0001** | 0.918 | 0.080 |
| Mean PR(SD), ms | 206.3(94.9) | 211.9(107.6) | 0.399 | 211.1(95.1) | 208.7(106.2) | 0.702 | 0.344 | 0.617 |
| High P or F waves, n(%) | 5(5) | 11(11) | **0.034** | 6(6) | 10(10) | 0.248 | 0.739 | 0.739 |
| High R, n(%) | 52(52) | 54(54) | 0.637 | 45(45) | 54(54) | 0.078 | 0.108 | 1.00 |
| Deep S, n(%) | 19(19) | 20(20) | 0.763 | 22(22) | 23(23) | 0.782 | 0.467 | 0.467 |
| High T, n(%) | 61(60) | 54(54) | 0.127 | 54(54) | 48(48) | 0.083 | 0.178 | 0.289 |
| Small QRS, n(%) | 26(26) | 25(25) | 0.827 | 45(45) | 40(40) | 0.251 | **0.002** | **0.003** |

**Figure 4.**
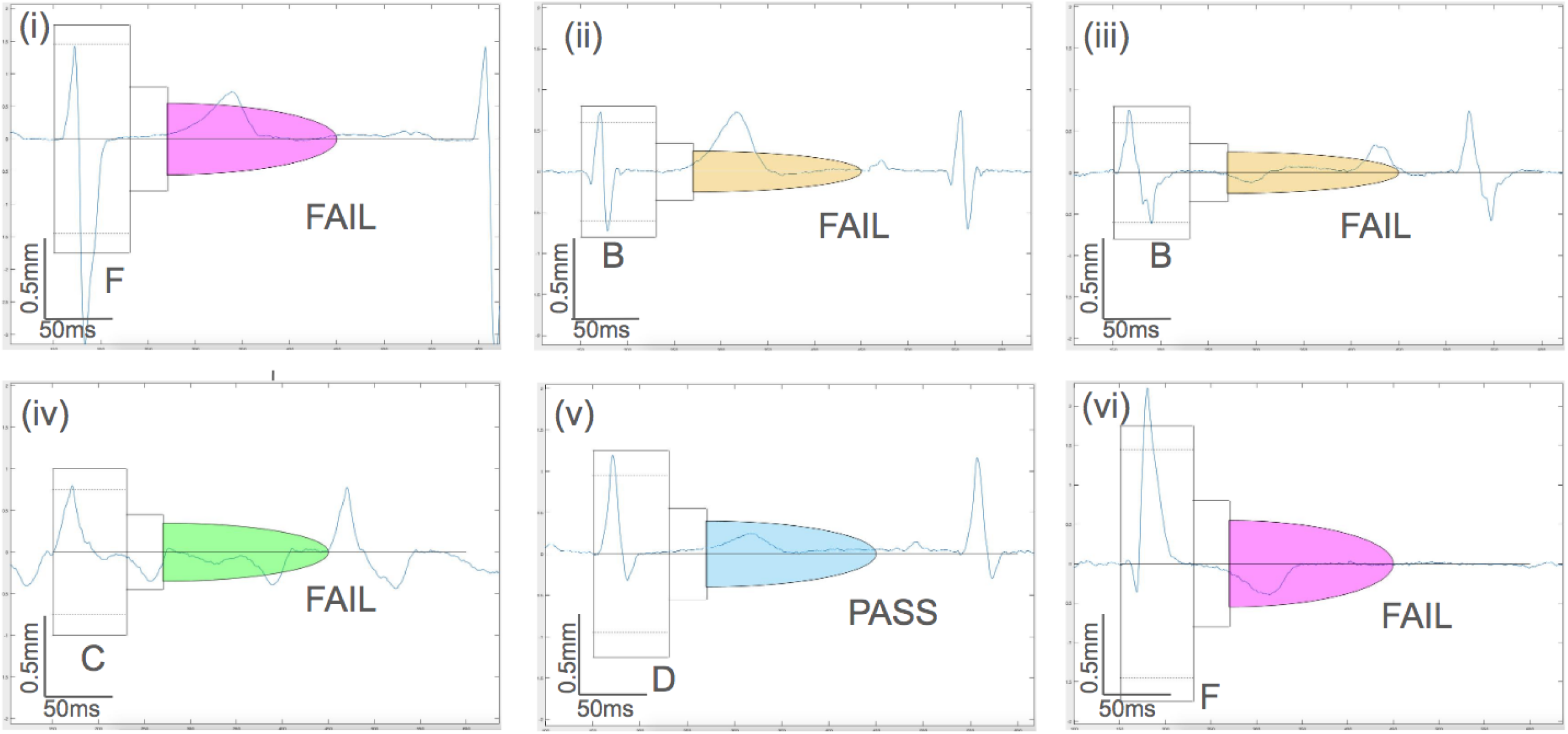
Representative examples of S-ICD screening template passing and failing ECG morphologies.

### Validation of the S-ICD eligibility prediction tool

Validation ROC AUC for our previous S-ICD eligibility tool^12^ was unsatisfactory (0.551; 95%CI 0.493 – 0.608). The sensitivity of the pre-defined threshold (≥ 0)^12^ was 73%, and specificity was 35%.

### Machine-learning prediction of S-ICD eligibility

Selection of the best models was performed using the supine ECG datasets. A comparison of the prediction models’ performance using testing supine ECG samples is shown in Table 3. For the left-sided S-ICD eligibility prediction, the ridge model demonstrated the smallest deviance, and the largest deviance ratio, which characterizes the best cross-validation function. The elastic net model was the 2^nd^ best, closely followed by lasso. Logistic regression showed the worst out-of-sample cross-validation function for both left-sided and right-sided prediction. Ridge and logistic regression models included all predictor variables, whereas lasso selected only four predictors (HR, QT interval, T_max_, and T_V1_ amplitude), and elastic net – only five predictors (HR, QT interval, T_max_, T_V1,_ and peak-to-peak QRS_V3_ amplitudes). Cross-validation plots and coefficient paths are shown in Figure 5. Using the lasso and elastic net prediction model estimates, left-sided S-ICD eligibility can be calculated as the following:

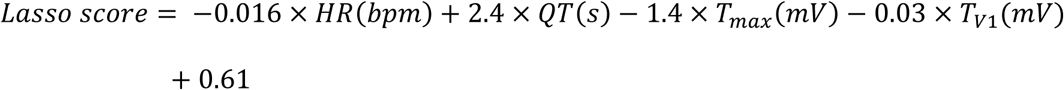

**Table 3.**
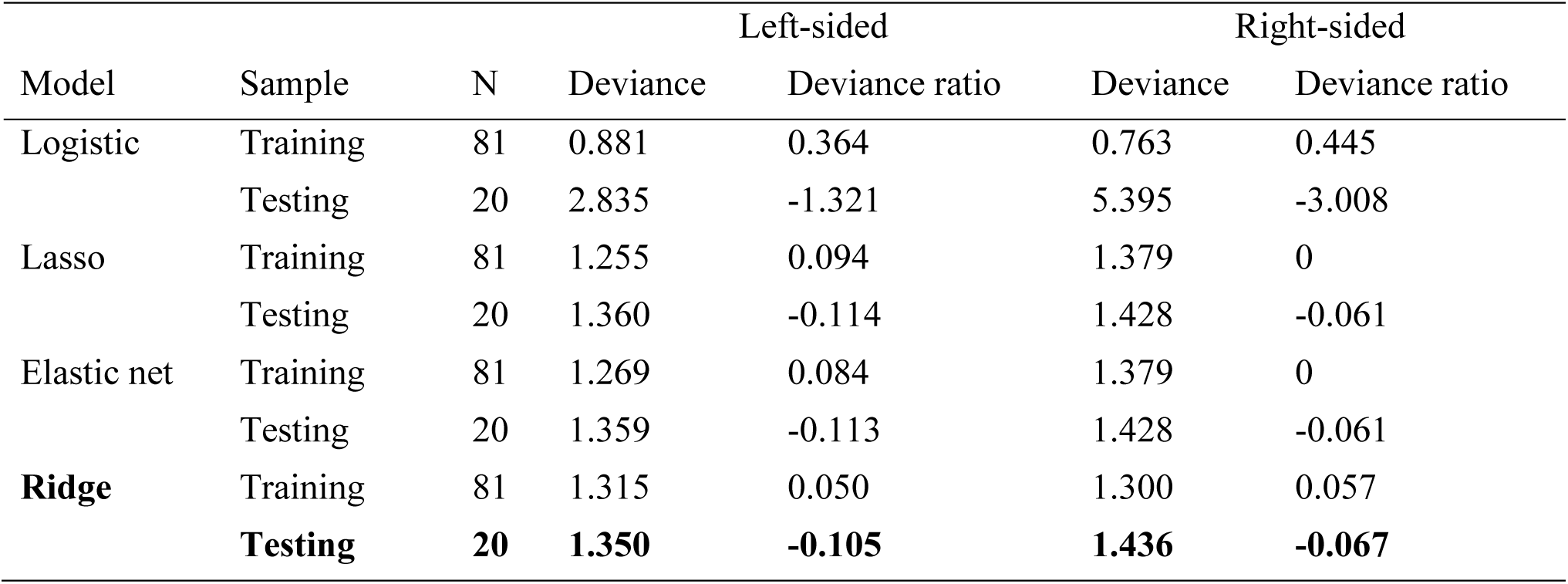
Machine learning model selection using supine ECG datasets

**Figure 5.**
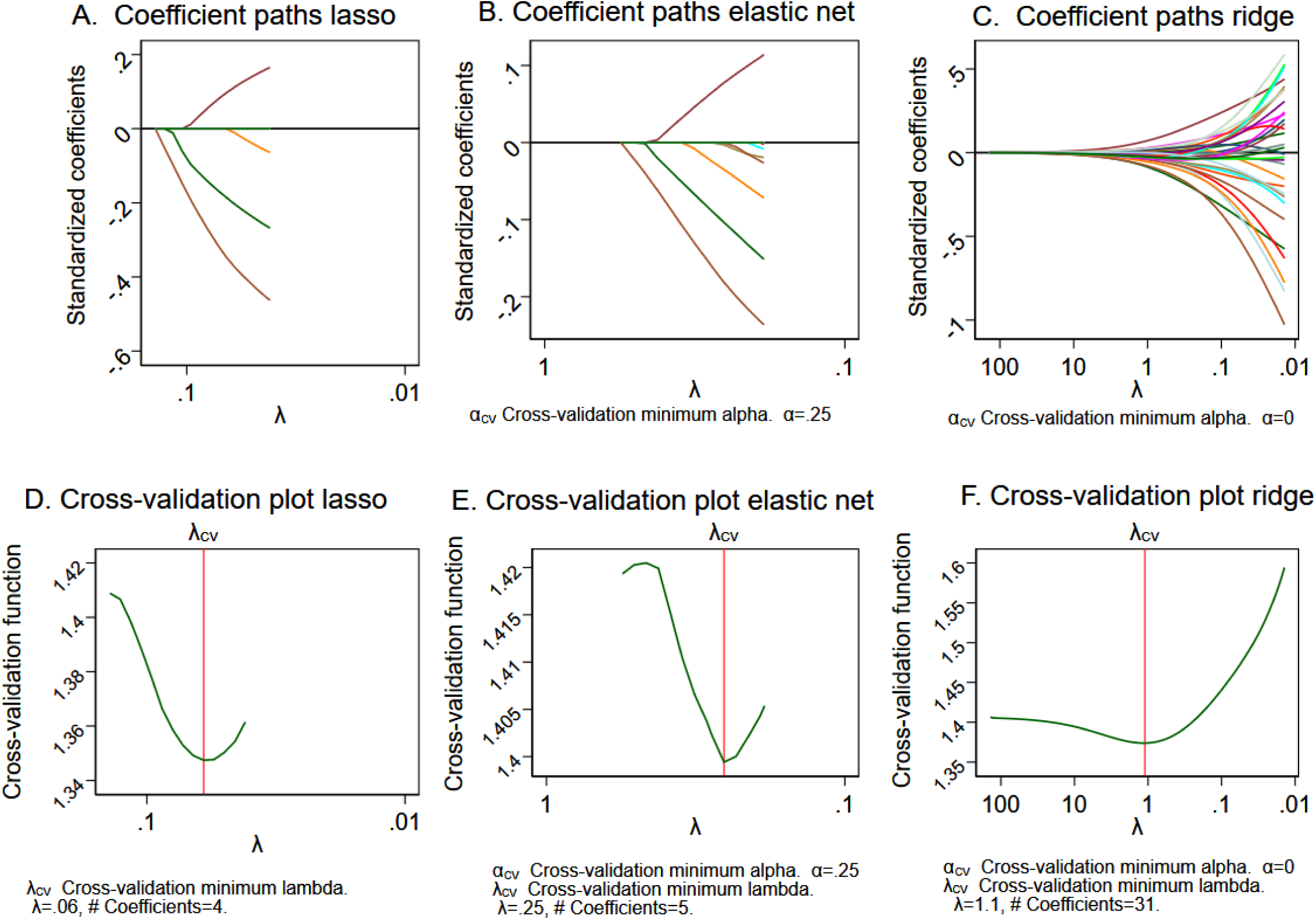
The coefficient paths after (**A**) lasso, (**B**) elastic net, (**C**) ridge models. A line is drawn for each coefficient that traces its value over the searched values of the lasso penalty parameter λ on a reverse logarithmic scale. Lasso is letting variables into the model based on its penalty and the current value of λ. Cross-validation (CV) function (the mean deviance in the CV samples) is plotted over the search grid for the lasso penalty parameter λ on a reverse logarithmic scale for (**D**) lasso, (**E**) elastic net, (**F**) ridge models. The first λ tried is on the left, and the last λ tried is on the right.

Lasso score ≥ - 0.5 predicted left-sided S-ICD eligibility with 91% sensitivity and 30% specificity.

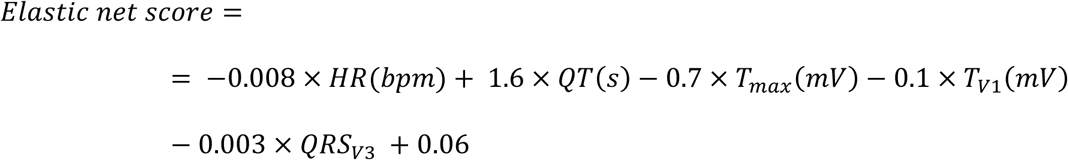

Elastic net score ≥ - 0.5 predicted left-sided S-ICD eligibility with 96% sensitivity and 10% specificity.

For the right-sided S-ICD eligibility prediction, both lasso and elastic net models shrunk to zero coefficients. Therefore, even if both lasso and elastic net demonstrated the minimum cross-validation function, we had to select the ridge model as the best model (Table 3). Therefore, we were not able to develop simple linear equations for right-sided S-ICD eligibility prediction.

Out-of-sample (standing ECG) validation of the ridge models was satisfactory for both left-sided [ROC AUC 0.687 (95%CI 0.582-0.791)] and right-sided [ROC AUC 0.655(95%CI 0.549-0.762)] S-ICD eligibility prediction.

Out-of-sample validation of the lasso and elastic net prediction models in the previous non-ACHD study population ^12^ yielded high sensitivity of the pre-selected in this study threshold (≥ - 0.5): 100% for the elastic net model, and 77% for lasso model. Validation ROC AUC in a non-ACHD population was unsatisfactory for all models: specifically for lasso (ROC AUC 0.554; 95%CI 0.355-0.754), elastic net (ROC AUC 0.548; 95%CI 0.340-0.756), and ridge model (ROC AUC 0.477; 95%CI 0.282-0.671).

### Transformation of routine clinical 12-lead ECG to S-ICD 3-lead ECG, and vice versa

Transformation matrices are reported (Supplemental Tables 1–2). Validation of the transformation matrices showed satisfactory agreement between the originally recorded and transformed signals (Figure 6 and Supplemental Table 3). For most of the leads (52/60; 87%), the difference in the voltage was not clinically meaningful (less than 0.1 mV). We provided open-source software application for transformations at https://github.com/Tereshchenkolab/S-ICD_eligibility.

**Figure 6.**
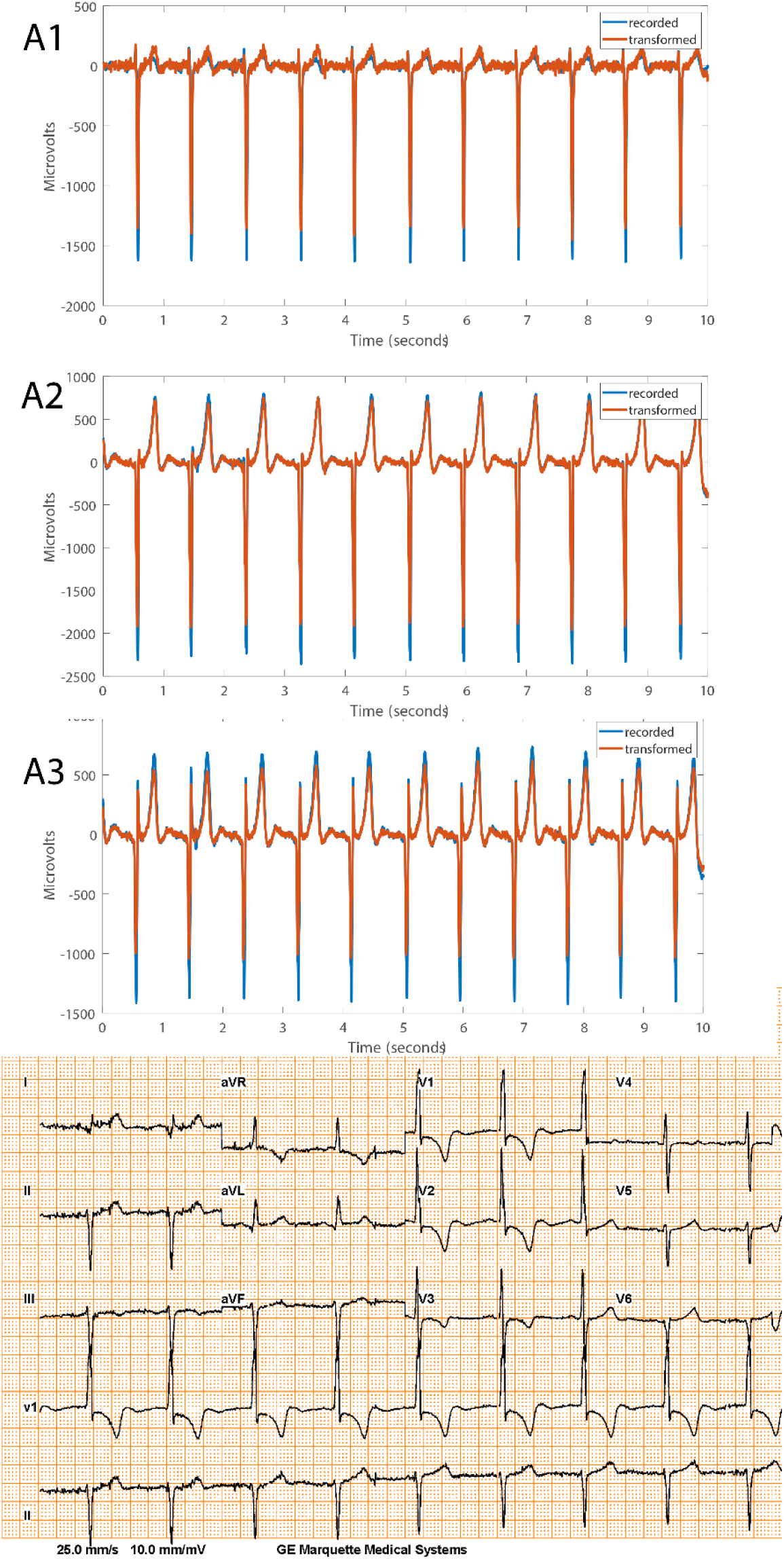
Representative examples of recorded and transformed right-sided 3-lead ECG morphologies, and corresponding 12-lead ECG recorded during standing in a Fontan patient.

## Discussion

This prospective study of the contemporary ACHD population revealed several important findings. First, we observed a high rate of S-ICD ineligibility: nearly half of ACHD patients were not eligible for S-ICD. While the complexity of ACHD was not associated with S-ICD ineligibility, ineligible ACHD patients exhibited a trend towards more significant hemodynamic impairment as compared to those who passed eligibility screening. The high rate of S-ICD ineligibility in ACHD population represents a significant barrier for the adoption of potentially advantageous and less invasive S-ICD technology for the prevention of sudden cardiac death in ACHD. Second, we used machine learning to develop and validate an S-ICD eligibility prediction tool, to simplify and make it more convenient to screen potential S-ICD candidates. We found that the most accurate prediction model suggests the use of as many as possible available 12-lead ECG features, and, therefore, is impractical for “by-hand” calculation.

Nevertheless, we were able to develop and validate a simplified S-ICD prediction model for left-sided S-ICD. The simplified model includes only four or five readily available ECG features; it has high sensitivity but low specificity and can be used as a first preliminary step for S-ICD eligibility screening. All calculators are freely available at www.ecgpredictscd.org. Thirdly, we developed and validated transformation matrices, to transform 12-lead ECG into S-ICD 3-lead ECG, and vice versa. The ability to reliably transform signals of these two leads systems could improve S-ICD diagnostics and facilitate the development of fully automated S-ICD eligibility assessment, using routinely recorded 12-lead ECG.

### Nearly half of the contemporary ACHD population is ineligible for S-ICD

In recent decades, the ACHD population has expanded due to the advancements in pediatric cardiology and congenital cardiac surgery; 90% of children with severe congenital heart disease now survive to 18 years of age.^14^ More than 1.4 million adults live with ACHD in the United States.^18^ Sudden cardiac death is the most frequent cause of death in ACHD.^19^ Patients with transposition of great arteries and tetralogy of Fallot have the highest risk of life-threatening ventricular arrhythmias.^20^ Since the entire S-ICD system is implanted in an extra-thoracic space, it eliminates the complications related to endo- or epicardial leads.^21^ The ACHD patients with no transvenous access to the heart (namely Fontal palliation), or those with a right-to-left shunt and increased risk of systemic emboli, can attain the utmost potential benefit^22^ from implantation of S-ICD. Unfortunately, our study demonstrated that 40% of the contemporary complex ACHD population is ineligible for S-ICD.

The rate of ineligibility observed in this study for both right- and left-sided S-ICD in ACHD patients (40%) is higher than the rate reported by Alonso et al.^23^ for tetralogy of Fallot (23%) and mixed ACHD patients^13^ (25%), the rate reported by Okamura et al. (12%)^24^, Garside et al.^6^ (25%; left-sided only), and Zeb et al.^25^ (13%; left-sided only). Higher rate of S-ICD ineligibility in our study can be due to the large size, greater complexity and heterogeneity, and more severe functional impairment of our study population.^14^ We found that the sickest patients with a trend towards higher degree of hemodynamic impairment (who potentially can benefit the most from S-ICD) have higher likelihood of failing ECG screening. The results of this study underscore the need to improve S-ICD technology further to increase the number of eligible ACHD patients. Our previous study^12^ showed remarkable (3-fold) improvement in S-ICD eligibility after ECG filtering. In this study, high QRS and T voltage was the main reason for S-ICD ineligibility and turning S-ICD ECG filtering feature ON can increase the number of eligible ACHD patients.

Similar to previous studies conducted in the ACHD population^13 23-25^, we found more Fontan and tetralogy of Fallot participants that passed screening with the right-sided vector. Findings of improved S-ICD eligibility with the right-sided placement of S-ICD lead merit further studies comparing effectiveness in arrhythmia termination. Several case reports demonstrated successful defibrillation with 65J in ACHD patients with right-sided S-ICD lead placement.^26–28^ Theoretically, right-sided S-ICD lead placement can be more effective in arrhythmia termination than left-sided S-ICD lead placement, because of a more favorable S-ICD electric lead field, encompassing the whole heart (Figure 1). An *in silico* study reported a lower defibrillation threshold for right-sided than for left-sided S-ICD lead placement.^29^ An observational study in a general S-ICD population^30^ demonstrated similar rates of successful defibrillation with the first 65J shock (79% left-sided and 73% right-sided lead; P=NS), and similar rates of ineffective shocks (2.9% left-sided and 1.9% right-sided lead; P=NS). A randomized controlled trial is needed before right-sided S-ICD lead placement can be recommended as preferential in ACHD.

### Using 12-lead ECG for prediction of 12-lead eligibility: a machine learning approach

Results of our study, demonstrating a large proportion of ACHD population being ineligible for S-ICD, highlight the importance of S-ICD eligibility screening. Currently, S-ICD eligibility assessment is performed in specialized centers, and some patients have to travel long distances only to be ultimately disqualified. The referring physician must assess S-ICD eligibility before offering this treatment option to a patient, to avoid disappointment if a patient is subsequently deemed ineligible. To address this constraint, we developed and validated S-ICD eligibility prediction tools, which can use widely available routine resting 12-lead ECG.

We used an advanced machine learning approach that illuminated several important findings. Model selection by machine learning demonstrated that the most accurate out-of-sample prediction tool included all available ECG features, specifically QRS and T amplitudes in each of 12 leads, all averaged ECG intervals (PR, QRS, QT), and heart rate. Along those lines, we developed transformation matrices to transform the entire ECG waveform from one type to another: from 12-lead ECG to 3-lead ECG and vice versa. Validation of transformation matrices demonstrated substantial agreement between originally recorded and transformed signals. Reliable signal transformation opens an avenue for further development of additional diagnostic and prognostic features that can enhance S-ICD functionality, as well as for the development of fully automated S-ICD eligibility assessment using routine 12-lead ECG.

At the same time, machine learning indicated that simplified prediction of S-ICD eligibility could not be sufficiently accurate. Both lasso and elastic net models for right-sided lead eligibility prediction shrunk all coefficients to zero, suggesting that no linear equation exist to describe prediction function accurately, because of its non-linearity. Therefore, we did not offer a simplified calculator to predict right-sided S-ICD eligibility. On the other hand, several models were selected by machine learning for the simplified prediction of left-sided S-ICD eligibility. Selected by machine learning S-ICD eligibility predictors (HR, QT, maximum T amplitude) have been previously reported in other ACHD studies^24^, including our previous model.^12^ Realizing that even using a machine learning approach we cannot offer perfect prediction of S-ICD eligibility by a simple linear model, we tuned developed models to high sensitivity. A simple calculator using readily available ECG metrics (HR, QT, T_max_, T_V1_, QRS_V3_) can be used for screening; it can identify all potential S-ICD candidates that need to undergo further assessment by the Boston Scientific EMBLEM S-ICD Patient Screening tool.^8^

Importantly, in this study, we used a supervised machine learning approach, to be able to interpret the models and understand factors associated with S-ICD ineligibility, and to provide open-source prediction tools. We cannot rule out a possibility of more accurate prediction by unsupervised machine learning, which was not utilized in this study.

### Limitations

We only performed ECG screening in the supine and standing positions but did not screen during exercise or other postures, which can theoretically increase the percentage of ineligible patients. Nonetheless, as we observed very little difference in eligibility between standing and supine positions in this study, we can infer that unlike in the general population,^12^ body posture change in an ACHD population has little to no effect on S-ICD eligibility. Consistently with our findings, Wilson et al.^31^ did not detect significant differences in the R/T amplitude ratio in tetralogy of Fallot and single ventricle physiology patients in a supine, prone, left lateral, right lateral, sitting, and standing positions, whereas such differences were observed in controls. Similarly, Zeb et al.^25^ reported that posture change did not affect S-ICD eligibility in ACHD patients.

On the other hand, in our study, an increase in HR was associated with large P-waves as a cause of ineligibility, and overall, with less likelihood of passing the screening. As ACHD patients are prone to sinus tachycardia and supraventricular arrhythmias, future studies of S-ICD eligibility in ACHD during exercise are needed.

Although we enrolled a complex ACHD population and presented a comparable sample size^6, 13, 24^, our study suffered limitations typical for all ACHD studies.^14^ ACHD patients are heterogeneous: specific congenital lesions and repairs are rare. Each ACHD patient has unique anatomy and physiology. Larger studies in ACHD populations would better account for inherent heterogeneity. However, our study cohort was representative of the wide variety of ACHD patients typically seen across the range of both anatomic and physiologic spectra, and thus the findings are likely more generalizable than other studies focusing on single defects. It is noteworthy that our broad inclusion also encompasses patients who would require a transvenous ICD because of indications for pacing and who would not be considered for S-ICD.

While our study had an equal presentation of men and women, the study population was predominantly white. Future studies in ethnically diverse populations are needed. It is not clear what role, if any, race or ethnicity would have on S-ICD eligibility.

Finally, though we found some suggestions of poorer hemodynamics in patients who were ineligible for S-ICD, there was no sufficient statistical power to detect differences in LVEF; estimated power was 0.34. RV and systemic ventricle hemodynamic function was not systematically evaluated in this study. Larger studies are needed to validate our finding of a trend towards greater hemodynamic impairment in unsuitable for S-ICD ACHD patients.

## Data Availability

A MATLAB (MathWorks, Inc, Natick, MA) open-source code for ECG analyses and a user manual is provided at https://github.com/Tereshchenkolab/S-ICD_eligibility. Fully de-identified raw digital ECG signal data generated for this study are available at https://github.com/Tereshchenkolab/S-ICD_eligibility. All calculators are freely available at www.ecgpredictscd.org.

http://www.ecgpredictscd.org/sicd-eligibility

## Acknowledgments

The authors thank the study participants and staff. We thank Christopher Hamilton, BA, and Meghan Hisatomi Saito, ACNP, for help with ECG recording and enrollment.

---

**Supplemental Table 1.**
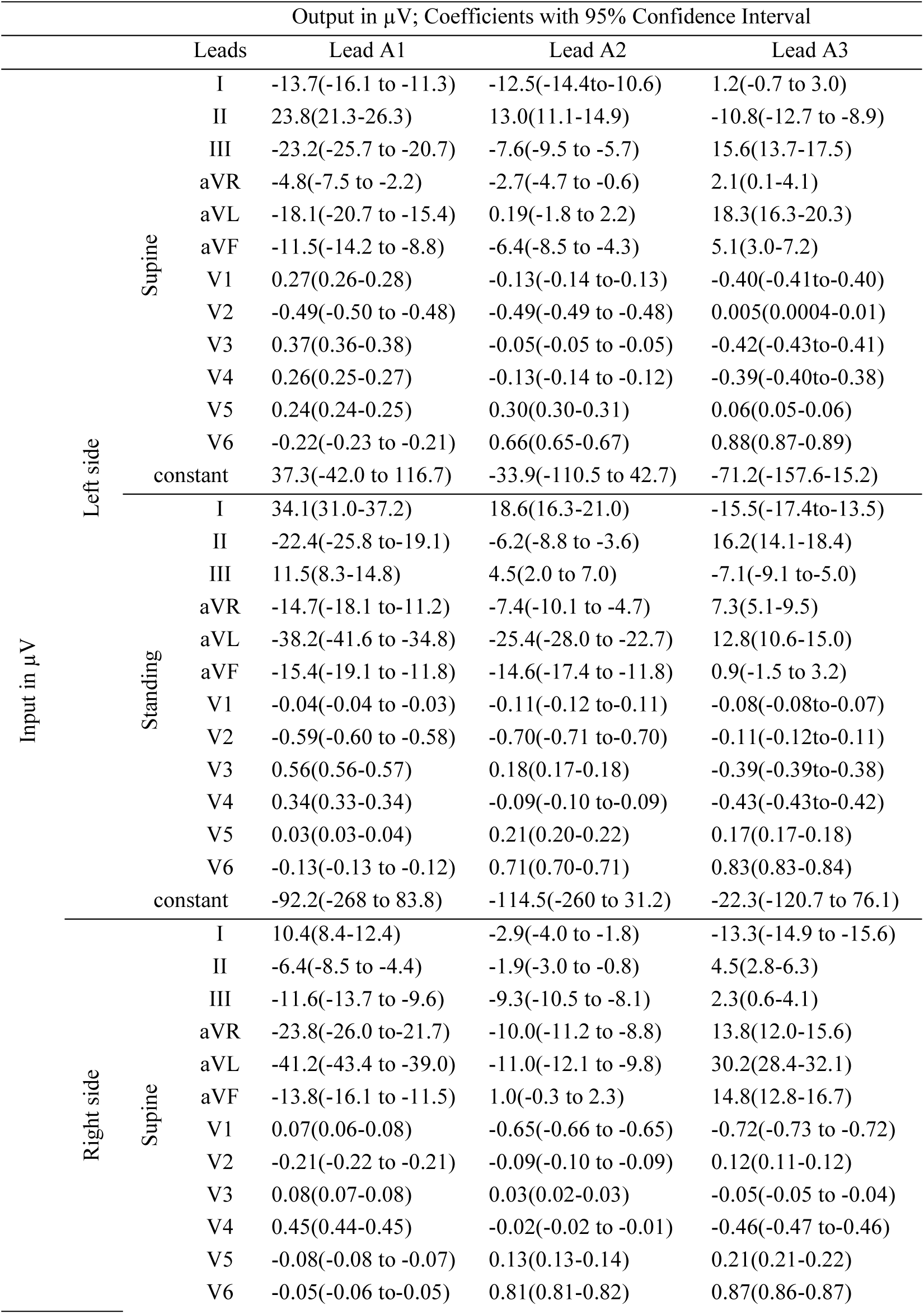
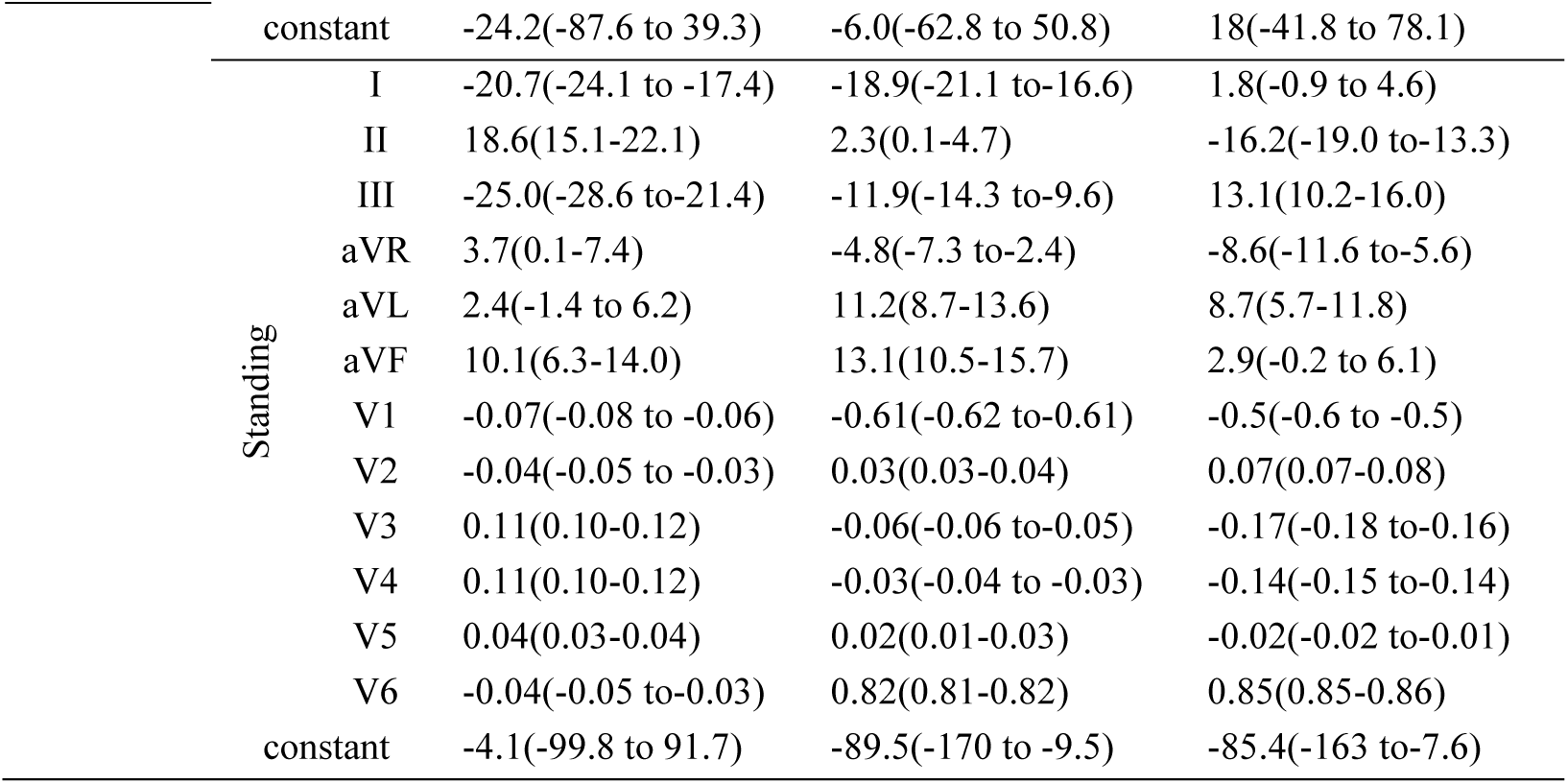
Transformation matrices from 12-lead ECG to 3-leas S-ICD ECG

**Supplemental Table 2.**
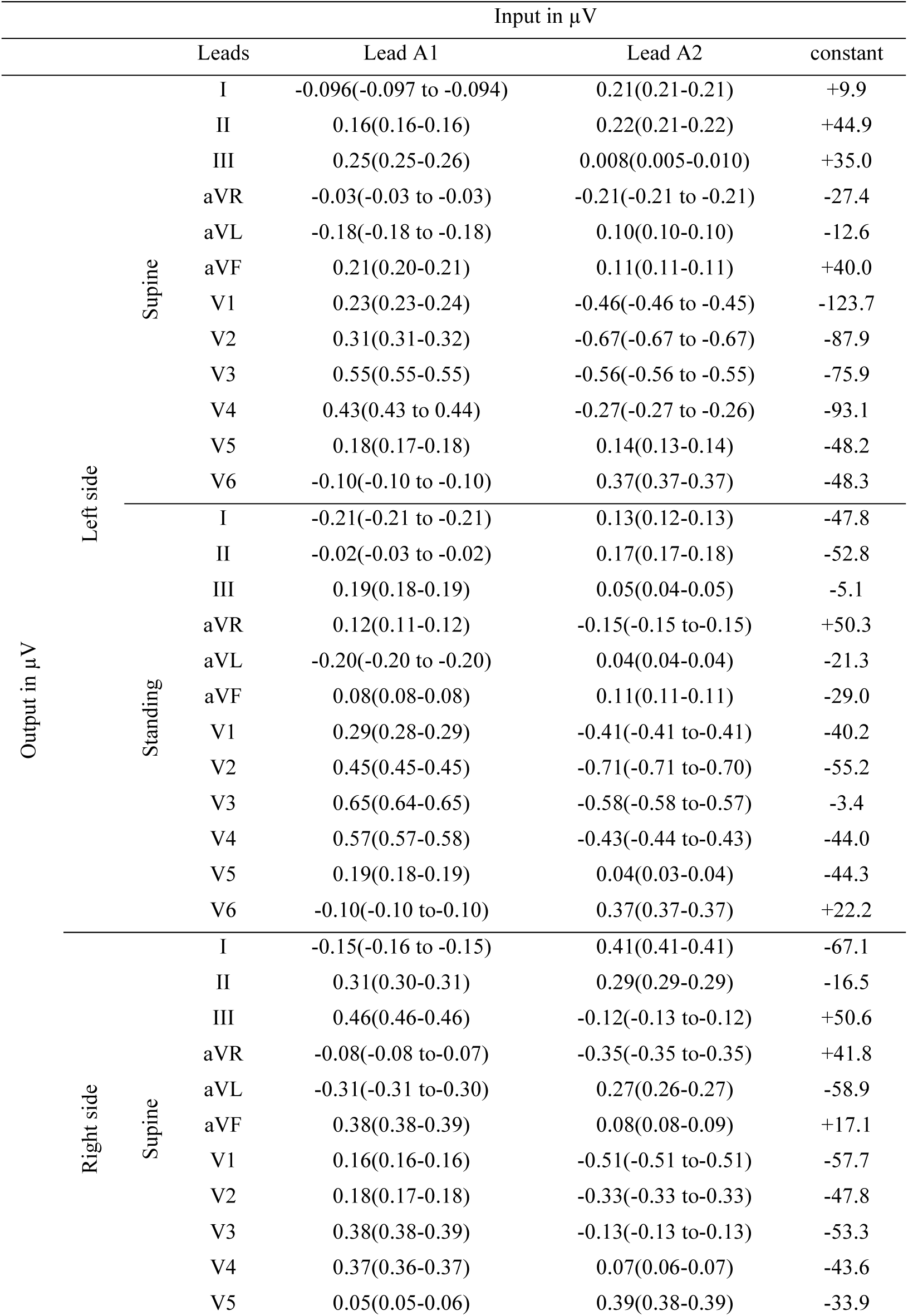
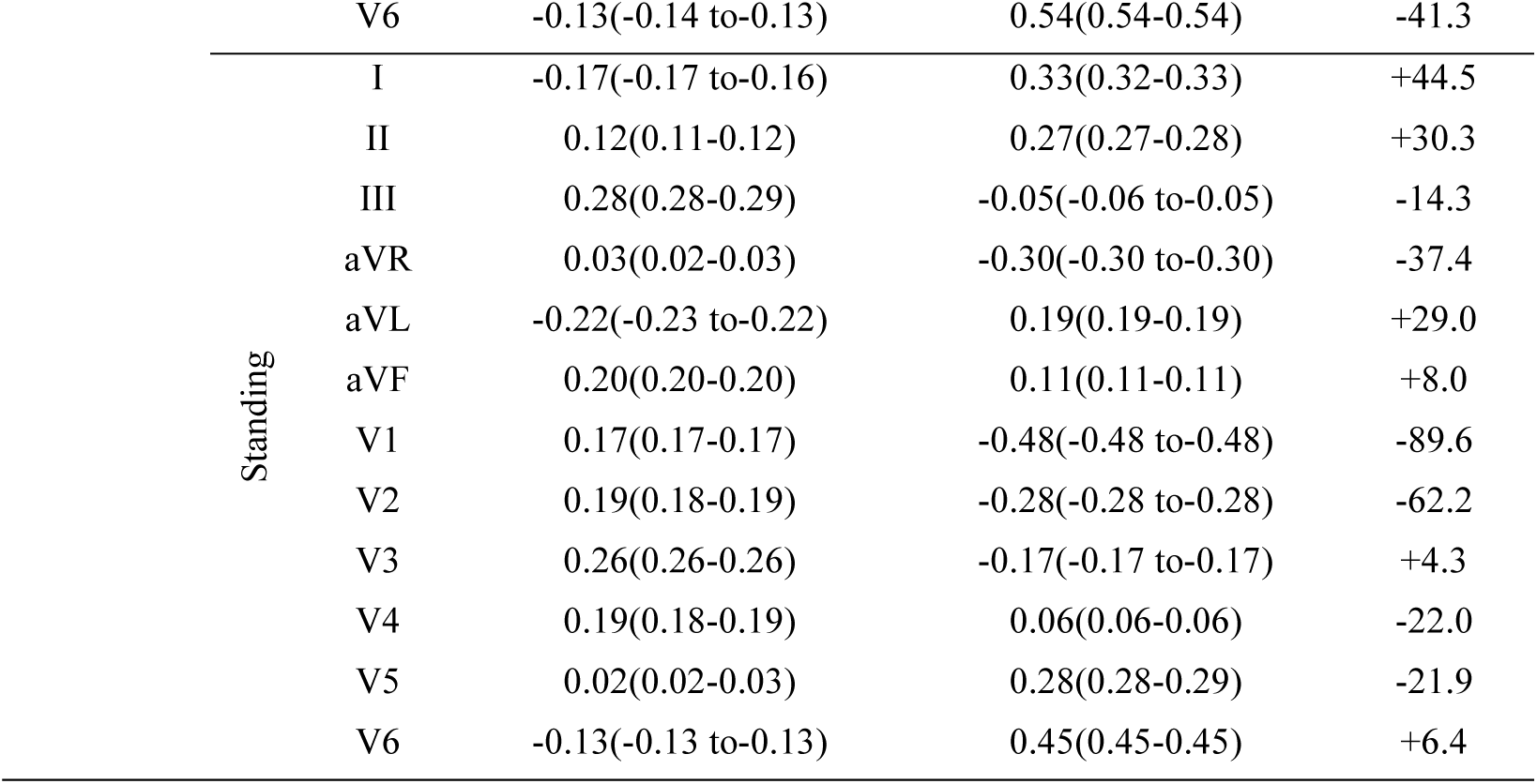
Transformation matrices from 3-leas S-ICD ECG to 12-lead ECG

**Supplemental Table 3.**
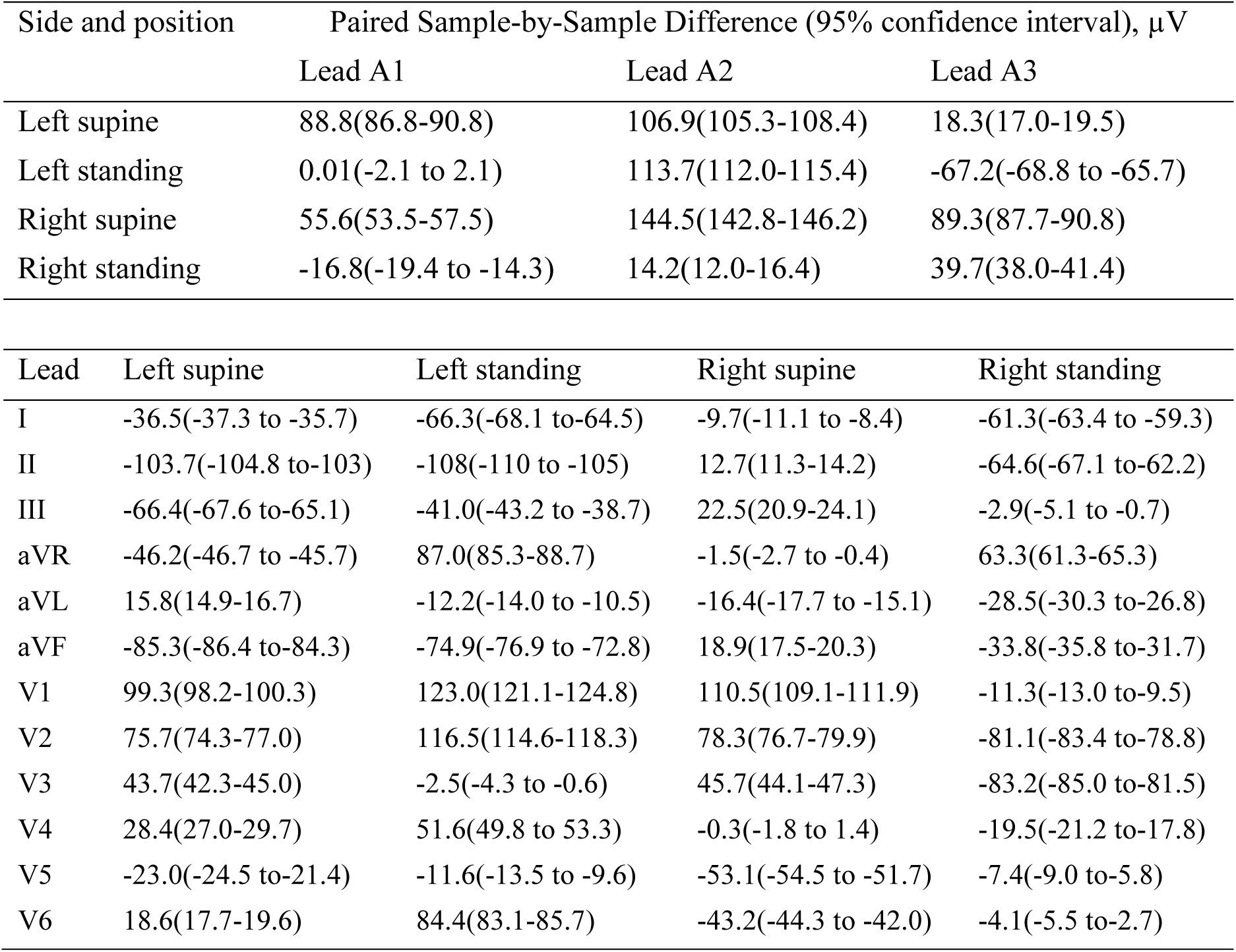
Validation of 12-lead to 3-lead S-ICD ECG, and vice versa transformation matrices

| Side and position | | Paired Sample-by-Sample Difference (95% confidence interval), $\mu\text{V}$ | | |
| --- | --- | --- | --- | --- |
|  |  | Lead A1 | Lead A2 | Lead A3 |
| Left supine |  | 88.8(86.8-90.8) | 106.9(105.3-108.4) | 18.3(17.0-19.5) |
| Left standing |  | 0.01(-2.1 to 2.1) | 113.7(112.0-115.4) | -67.2(-68.8 to -65.7) |
| Right supine |  | 55.6(53.5-57.5) | 144.5(142.8-146.2) | 89.3(87.7-90.8) |
| Right standing |  | -16.8(-19.4 to -14.3) | 14.2(12.0-16.4) | 39.7(38.0-41.4) |

| Lead | Left supine | Left standing | Right supine | Right standing |
| --- | --- | --- | --- | --- |
| I | -36.5(-37.3 to -35.7) | -66.3(-68.1 to -64.5) | -9.7(-11.1 to -8.4) | -61.3(-63.4 to -59.3) |
| II | -103.7(-104.8 to -103) | -108(-110 to -105) | 12.7(11.3-14.2) | -64.6(-67.1 to -62.2) |
| III | -66.4(-67.6 to -65.1) | -41.0(-43.2 to -38.7) | 22.5(20.9-24.1) | -2.9(-5.1 to -0.7) |
| aVR | -46.2(-46.7 to -45.7) | 87.0(85.3-88.7) | -1.5(-2.7 to -0.4) | 63.3(61.3-65.3) |
| aVL | 15.8(14.9-16.7) | -12.2(-14.0 to -10.5) | -16.4(-17.7 to -15.1) | -28.5(-30.3 to -26.8) |
| aVF | -85.3(-86.4 to -84.3) | -74.9(-76.9 to -72.8) | 18.9(17.5-20.3) | -33.8(-35.8 to -31.7) |
| V1 | 99.3(98.2-100.3) | 123.0(121.1-124.8) | 110.5(109.1-111.9) | -11.3(-13.0 to -9.5) |
| V2 | 75.7(74.3-77.0) | 116.5(114.6-118.3) | 78.3(76.7-79.9) | -81.1(-83.4 to -78.8) |
| V3 | 43.7(42.3-45.0) | -2.5(-4.3 to -0.6) | 45.7(44.1-47.3) | -83.2(-85.0 to -81.5) |
| V4 | 28.4(27.0-29.7) | 51.6(49.8 to 53.3) | -0.3(-1.8 to 1.4) | -19.5(-21.2 to -17.8) |
| V5 | -23.0(-24.5 to -21.4) | -11.6(-13.5 to -9.6) | -53.1(-54.5 to -51.7) | -7.4(-9.0 to -5.8) |
| V6 | 18.6(17.7-19.6) | 84.4(83.1-85.7) | -43.2(-44.3 to -42.0) | -4.1(-5.5 to -2.7) |

**Supplemental Figure 1:**
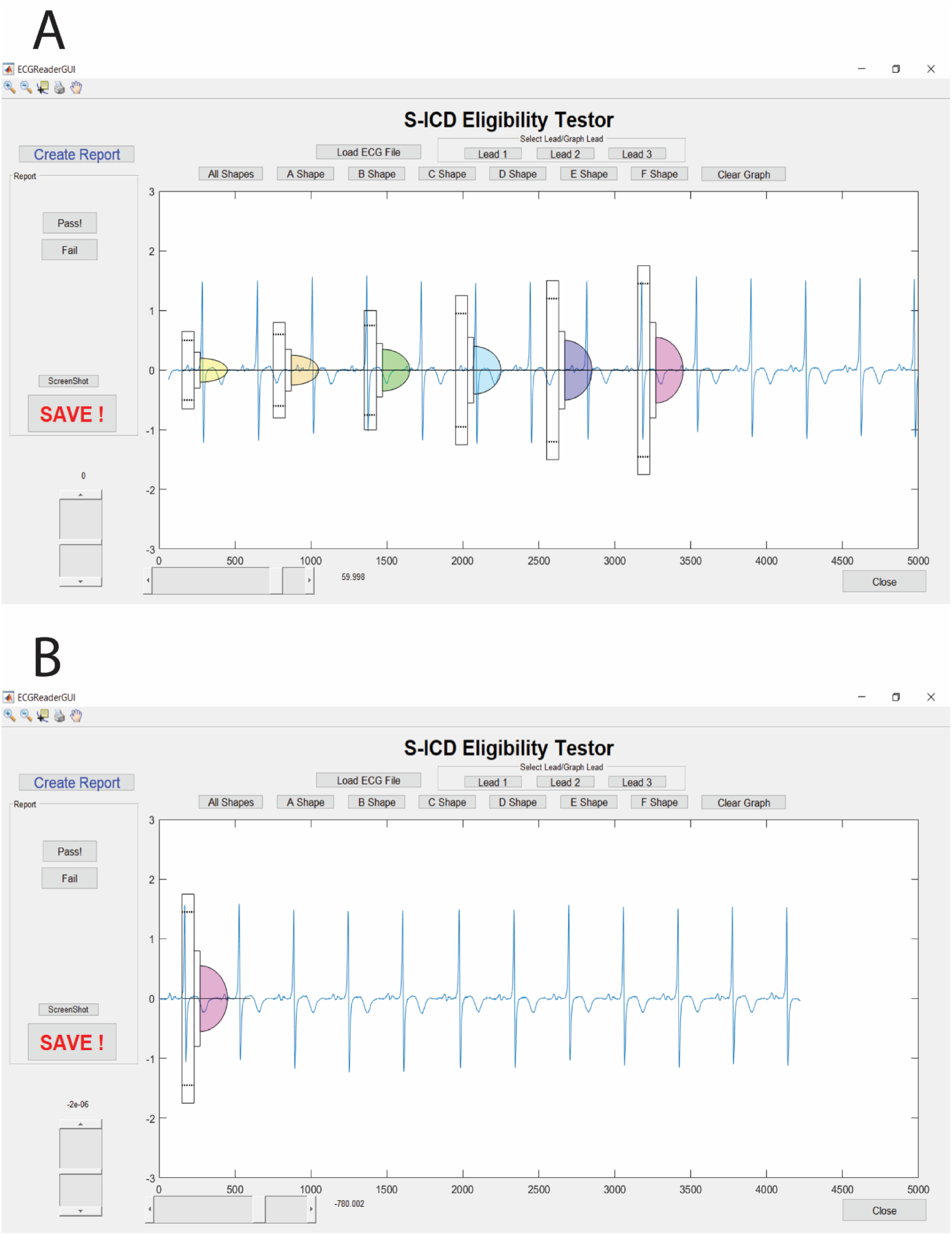
Layout of the S-ICD Eligibility Viewer. **A**. Select ‘Create Report’ button only once to record the data on text file. Load the 3-Lead ECG file by clicking the ‘Load ECG File’ button. Then select the appropriate lead to plot the ECG on the graph. Select a shape to start analyzing eligibility. **B**. Move the graph by using the slider at the bottom left corner of the screen. Use the zoom function on the upper left corner of screen to determine eligibility. Select ‘Pass’ or ‘Fail’ button to record eligibility. If ‘Fail’ button is clicked, select the corresponding reason for failure. Snap a screenshot of graph using ‘ScreenShot’ button. Click ‘SAVE’ button at the end of each lead to record information in the text file. All screenshots and the text file will be saved in same folder as the Viewer.

